# Determinants of COVID-19 outcomes: A systematic review

**DOI:** 10.1101/2021.03.21.21254068

**Authors:** Shirley Crankson, Subhash Pokhrel, Nana Kwame Anokye

**Affiliations:** Division of Global Public Health, Department of Health Sciences, College of Health, Medicine and Life Sciences, Brunel University London

## Abstract

**Background:** The current pandemic, COVID-19, caused by a novel coronavirus SARS-CoV-2, has claimed over a million lives worldwide in a year, warranting the need for more research into the wider determinants of COVID-19 outcomes to support evidence-based policies.

**Objective:** This study aimed to investigate what factors determined the mortality and length of hospitalisation in individuals with COVID-19.

**Data Source:** This is a systematic review with data from four electronic databases: Scopus, Google Scholar, CINAHL and Web of Science.

**Eligibility Criteria:** Studies were included in this review if they explored determinants of COVID-19 mortality or length of hospitalisation, were written in the English Language, and had available full-text.

**Study appraisal and data synthesis:** The authors assessed the quality of the included studies with the Newcastle□Ottawa Scale and the Agency for Healthcare Research and Quality checklist, depending on their study design. Risk of bias in the included studies was assessed with risk of bias assessment tool for non-randomised studies. A narrative synthesis of the evidence was carried out. The review methods were informed by the Joana Briggs Institute guideline for systematic reviews.

**Results:** The review included 22 studies from nine countries, with participants totalling 239,830. The included studies’ quality was moderate to high. The identified determinants were categorised into demographic, biological, socioeconomic and lifestyle risk factors, based on the Dahlgren and Whitehead determinant of health model. Increasing age (ORs 1.04-20.6, 95%CIs 1.01-22.68) was the common demographic determinant of COVID-19 mortality while living with diabetes (ORs 0.50-3.2, 95%CIs −0.2-0.74) was one of the most common biological determinants of COVID-19 length of hospitalisation.

**Review limitation:** Meta-analysis was not conducted because of included studies’ heterogeneity.

**Conclusion:** COVID-19 outcomes are predicted by multiple determinants, with increasing age and living with diabetes being the most common risk factors. Population-level policies that prioritise interventions for the elderly population and the people living with diabetes may help mitigate the outbreak’s impact.

**PROSPERO registration number:** CRD42021237063.

**Strength and limitations of this review:**

- This is the first systematic review synthesising the evidence on determinants of COVID-19 LOS outcome.
- It is also the first review to provide a comprehensive investigation of contextual determinants of COVID-19 outcomes, based on the determinants of health model; thus, presenting with crucial gaps in the literature on the determinants of COVID-19 outcomes that require urgent attention.
- The review was restricted in conducting meta-analysis due to included studies’ heterogeneity.
- The review focused on only papers published in the English Language; hence, other relevant papers written on other languages could have been omitted.

## Introduction

COVID-19 (SARS-CoV-2, coronavirus) is currently among the leading causes of death globally. As of January 9th, 2021, 87,589,206 cases and 1,906, 606 deaths had been recorded globally (1). While its case fatality ratio (CFR) has been relatively low (CFR=2.2%), compared to CFRs of previous coronavirus outbreaks, notably, MERS (CFR=9.5%) and SARS-COV-1 (CFR=34.4%), its aggressive and alarming transmission rate has posed enormous challenges on global health (2). Even the current reported transmission rate of

COVID-19 may be lower than the actual transmission rate because a significant proportion of infected persons may remain undetected because they are asymptomatic (3). And regarding its CFR, current predictions even suggest that mortality ratio may increase since the pandemic is still ongoing (4). The infectivity and fatality rates associated with COVID-19, together with the worldwide panic it generates, make the current coronavirus pandemic a significant threat to public health, and its gargantuan impact unlike anything the world has experienced in the last two decades (5).

Since its inception, COVID-19 has overburdened the whole global health system, from crippling health resources to causing paradigm shifts in health care delivery (6). The testing process, quarantine and isolation associated with the virus has had dire psychological and financial implications on individuals and institutions (7). Furthermore, lockdowns instituted by affected countries to curb the virus’s spread resulted in disrupted formal education, unplanned fiscal costs on emergency reliefs, and decreased productivity, all translating into huge economic costs to governments and organisations (8). The overall COVID-19 burden, in terms of health and fiscal implications, has been consequential in both high-income and low- and middle-income countries, albeit with contextual differences.

Regardless of the significant interventions to curb the virus’s spread and subsequently reduce its severest outcome, i.e., mortality and morbidity, the outbreak continue to increase. As of January 9th, 2021, the daily global COVID death was 15,522, the highest daily mortality since the pandemic started, and about 3,000 more deaths since the first peak in daily COVID-19 deaths (initial peak April 17th – 12,511 daily deaths) (1). Also, 823,856 cases were confirmed on January 9th, 2021, representing a 0.93% increase from the previous day’s case count. The rapid rise in the COVID-19 cases and deaths worldwide necessitates continuous research on risk factors for COVID-19 outcomes to provide current evidence-based interventions to reduce the outbreak’s drastic impact.

Several studies on determinants of COVID-19 outcomes were identified in the literature; however, most of them were primary studies investigating risk factors for COVID-19 mortality (9, 10, 11). The few secondary studies/systematic reviews found in the literature encompassed only papers from high-income countries (12, 13). Also, there is inadequate coverage on other COVID-19 outcomes, specifically, length of hospital stay (LOS). Till date, no review has explored risk factors/determinants of COVID-19 LOS. Findings from such studies will be essential for help health systems to develop contingency plans for bed occupancy and health resources, especially with the swift increase in the COVID-19 cases. Thus, this study aimed to review factors that determine COVID-19 mortality and LOS in individuals diagnosed with COVID-19 to address the literature dearth and contribute to global efforts at curtailing the pandemic. Understanding the risk factors of COVID-19 outcomes based on a comprehensive synthesis of global but rapidly emerging evidence might be useful to implement effective policies to address the disease burden.

## Methods

### Search strategy

From 21^st^ to 31^st^ December 2020, Scopus, Google Scholar, CINAHL and Web of Science databases were searched for relevant studies using the search terms: ‘Determinants’ ‘Predictors’ ‘COVID-19’ ‘SARS-CoV-2’ ‘Mortality’ ‘Length of hospital stay’ ‘Length of hospitalisation’. The search terms were combined with mesh words and Boolean operators to ensure sensitive and targeted search. Full search strategy is shown in supplementary information 1. First screening of the databases results was conducted independently by two of the reviewers (SC and NKA) to ensure their relevance to this study. The tiles and abstract of the identified relevant studies were screened against this review’s eligibility using the following predetermined eligibility criteria: population - individuals diagnosed with COVID-19, exposures – demographic, socioeconomic, lifestyle, environmental biological/medical factors, outcome – COVID-19 related mortality and LOS, studies that explored determinants of COVID-19 mortality and LOS in participants with COVID-19, studies whose LOS endpoint was discharge or death, and not hospital transfers, studies written and published in the English Language, and with full-text available. No date restriction was applied in any of the databases since most COVID-19 studies are recent. No database filters were also applied. The references of the identified papers were also tracked for papers eligible for this review. Any disagreements relating to studies’ screening was discussed and resolved with the third reviewer (SK) and

### Data extraction

Two of the authors (SC and NKA) extracted the relevant data from the included studies using a comprehensive a priori developed set of data extraction questions (Supplementary information 2), informed by the JBI data extraction tools. The questions were categorised under two main themes, i.e., general information (authors name, study settings, study aim, year of publication) and methodology (sample size, sample characteristics, outcomes etc.), to ensure the sensitivity of the questions to the overarching objectives of this review. The data extraction questions were piloted-tested on five selected studies before their final usage. The third author (SP) randomly selected and reviewed 50% of the extracted data from the included studies to ascertain data extraction quality. Finally, all disagreements were resolved by consensus.

### Risk of bias and quality assessment

The Newcastle□Ottawa Scale (NOS) and the Agency for Healthcare Research and Quality (AHRQ) appraisal checklist were used to appraise the quality of the included studies. These checklists were based on a systematic review’s recommendation (14). The NOS provides eight items grouped under three main domains: the selection of cohorts, the comparability of cohorts, and outcomes assessment. A star (*) was awarded if a study met an item under the three defined domains. A maximum of one star was given to items within the selection and outcome domain, and a maximum of two stars was given to the item under the comparability domain. Thus, studies with nine stars were rated as high-quality study and those with two stars or less were graded as low quality. Like the NOS, the ARHQ also provides items/checklists (n=11) for assessing the quality of the study’s’ methods and outcomes. A ‘yes’, ‘no’ or ‘not applicable (NA)’ was used to indicate whether a study met the AHRQ requirement. The number of ‘yes’ from a study represented the study’s quality. Consequently, studies with eleven ‘yes’, suggesting 11 total scores, were ranked as high quality, whereas those with two or less ‘yes’ were rated as low quality. Also, risk of bias assessment of both the study and outcome level of the included papers was performed with risk of bias assessment tool for non-randomised studies. The quality and risk assessment findings are presented in supplementary information 3.

### Data synthesis

Descriptive data synthesis, informed by the JBI manual for evidence synthesis in systematic reviews was conducted to comprehensively describe the methods, findings, and quality of the included studies (15). The studies’ methods, its operationalisation and the subsequent findings were compared across the papers to identify common determinants of COVID-19 mortality and LOS. Also, the range of effect sizes (odds ratios/hazard ratios) of the identified determinants in the studies were synthesised to understand the magnitude of the effect on the study outcomes. This was done by reporting the lowest and highest effect sizes from studies that identified common determinants.

### Patient and public involvement

This study reviewed already published and available research. Therefore, no patients or the public were directly involved in this review process. The findings of this review will be shared publicly through scientific publication, social media, and conference presentations.

## Results

### Search result

The database search yielded 1,653 studies. The authors removed 11 duplicates and eliminated a further 1,564 after title screening for relevance to this review. The abstracts of the remaining 78 studies were assessed for eligibility using the predetermined eligibility criteria. Twenty-two studies met the inclusion criteria and were subsequently included in this review, as shown in the Prisma flow diagram below (Fig 1).

**Fig 1.**
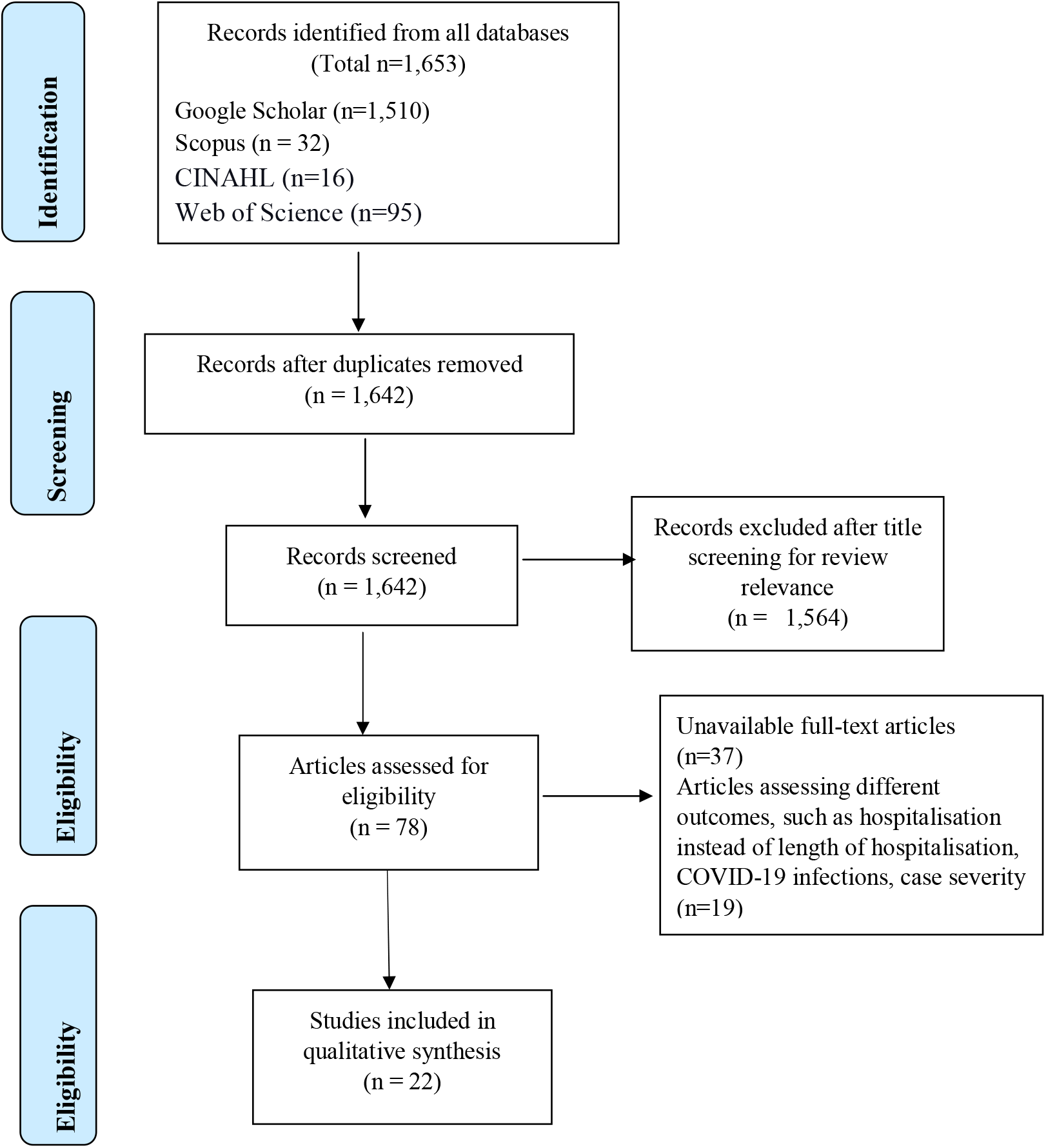
Prisma flow diagram showing the literature search.

### Overview of included studies

The studies spanned nine major countries, of which eight were high-income countries and one upper-middle-income country. Majority of them were from China (n=6) and USA (n=6), followed by Spain (n=2), and England (n=2), and one each from Kuwait, Mexico, France, Italy, Austria, and one multi-continent study-participants from Africa, Europe, Australia, Asia, and Americas (16). They all used the quantitative research approach, with retrospective cohort design as the predominant study design (n=12). Most of them (n=20) accessed only secondary data, retrieved mainly from the patients’ electronic medical records. The remaining two used both secondary and primary data (face to face and telephone interviews) (10, 17).

The studies sample size ranged from 58 to 177,133, totaling 239,830 participants. All the studies included both male and female participants; however, the men dominated, representing 51.4% of the studies’ entire population. Only 7 studies were age-specific - limited their inclusion to participants ≥18 years old. The majority (n=20) included only persons with confirmed COVID-19 test, either through the reverse transcription-polymerase chain reaction (RT-PCR) or nasopharyngeal swabs. The other two studies included all confirmed, negative, and suspected, and both suspected and confirmed COVID-19 cases (18, 19) (Table 1). The studies’ quality on the NOS and AHRQ ranged from 6-9, representing moderate to high quality and the common risk of bias across the studies was the inability to control the influence of unmeasured confounders. This confounding bias inherently influenced the studies’ findings and could subsequently affect this review’s finding.

**Table 1:**
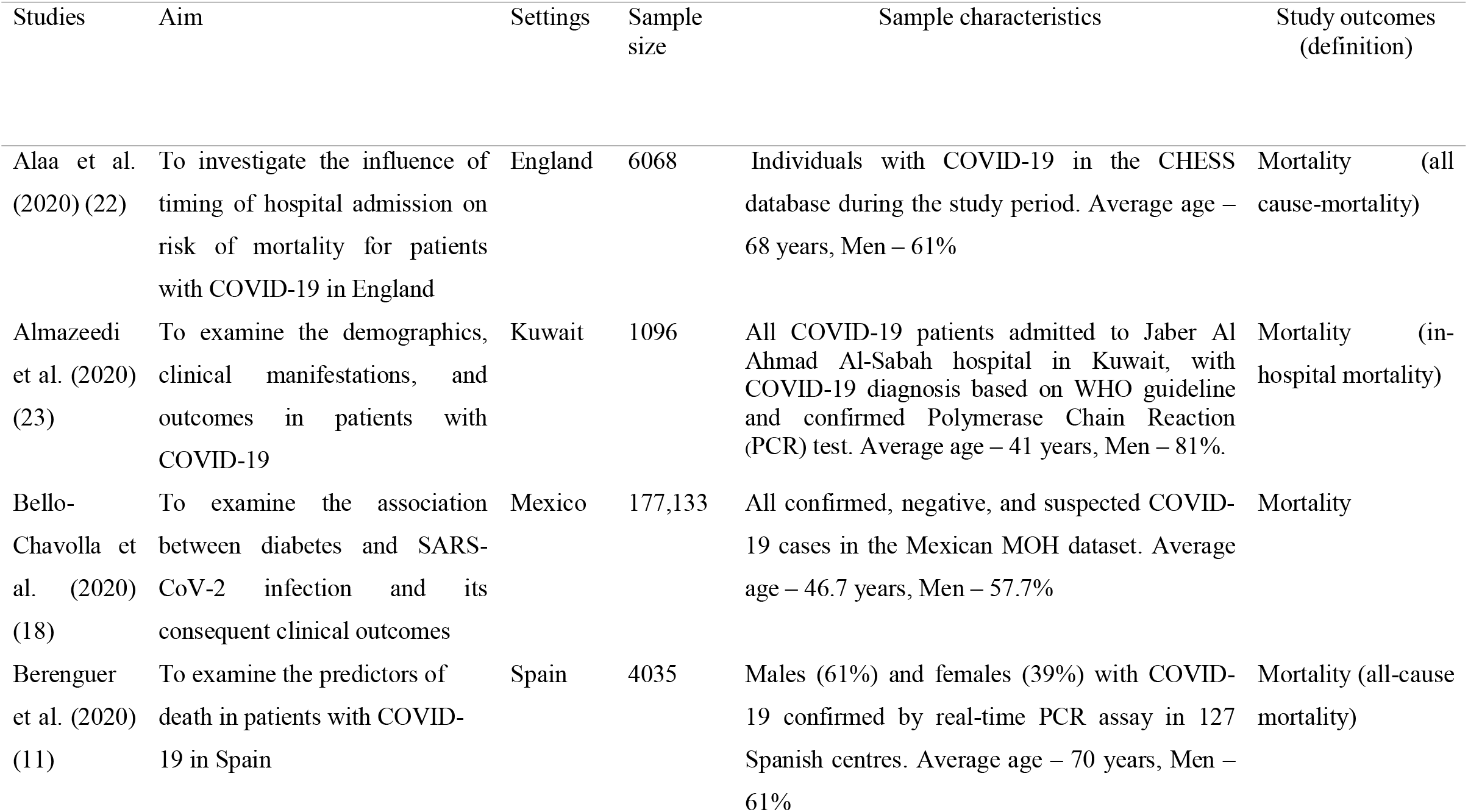
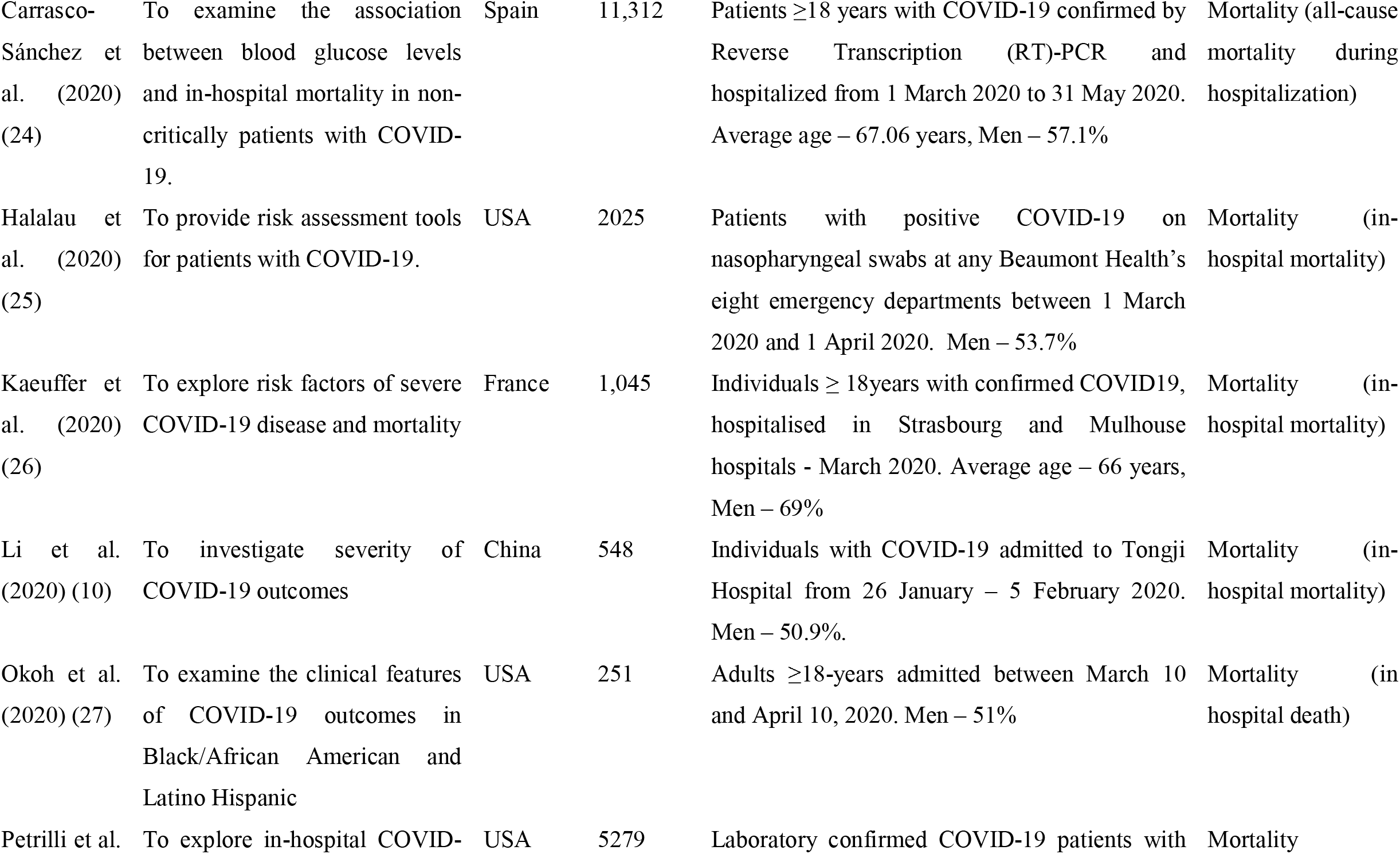
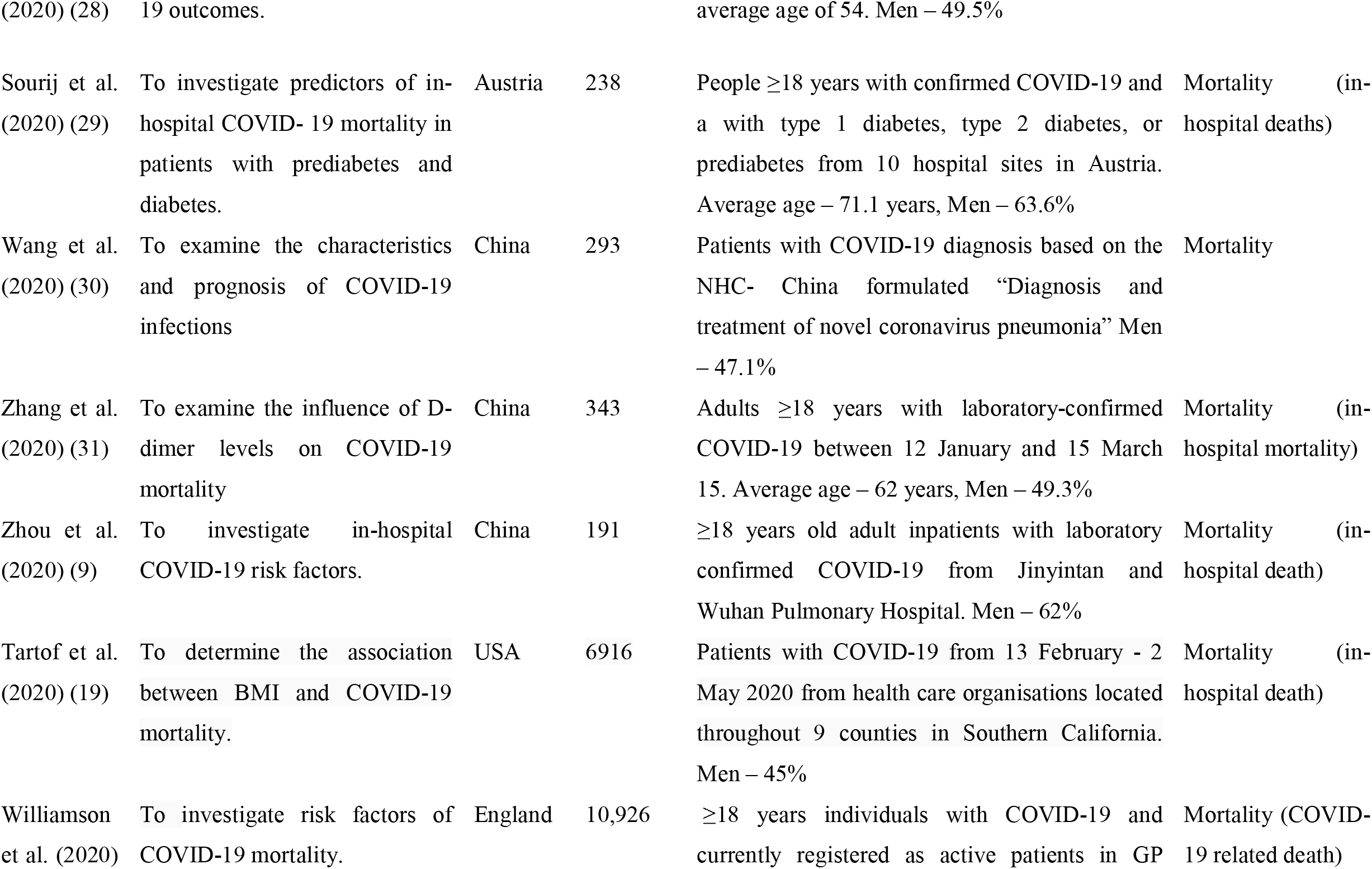
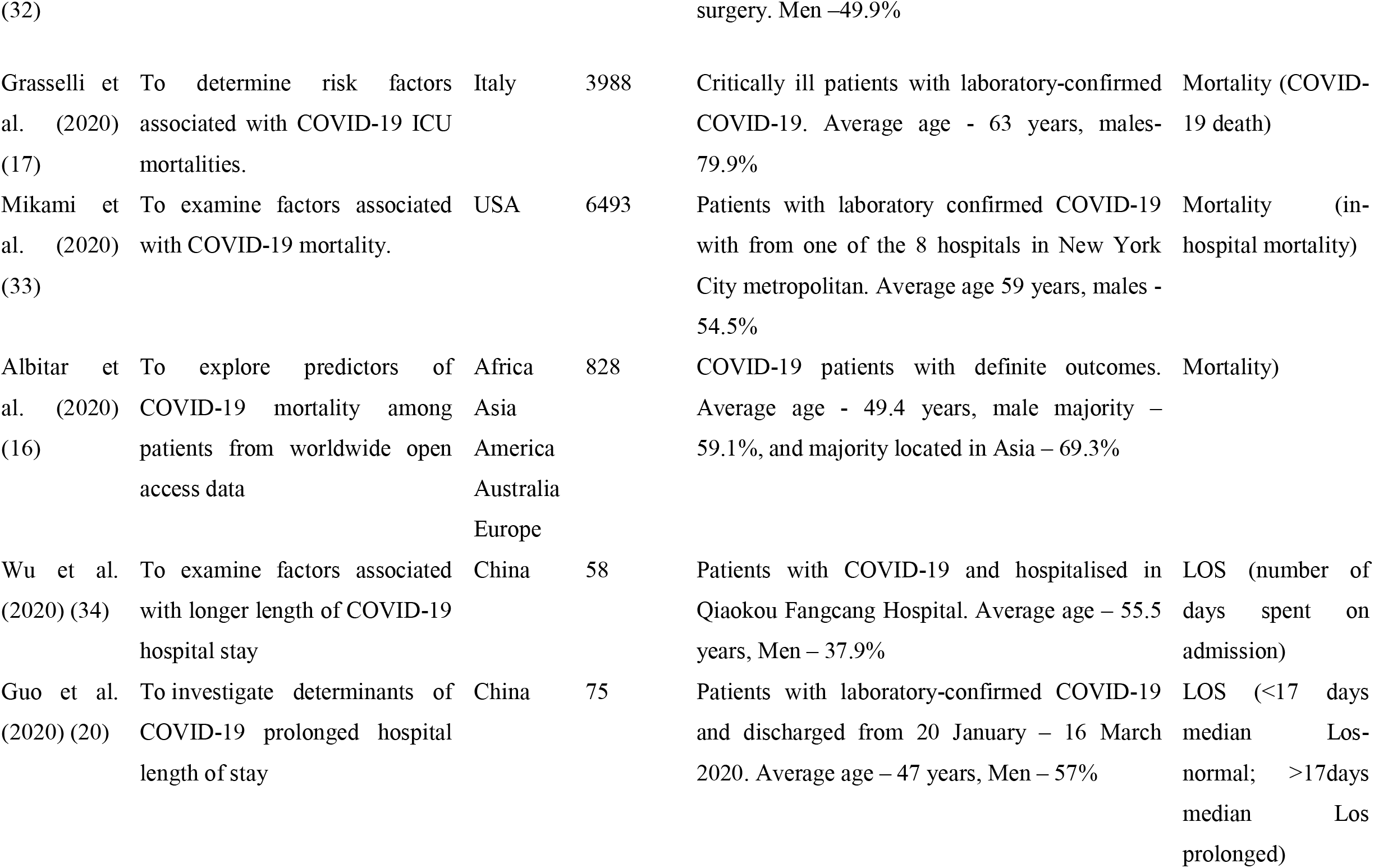
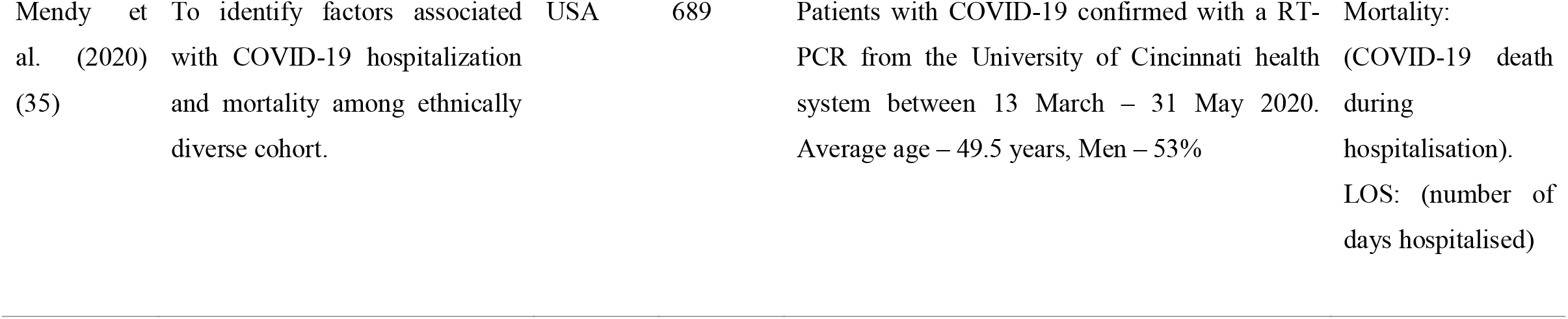
Study characteristics

Of the COVID-19 outcomes, 19 studies focused on only COVID-19 mortality, 2 solely on COVID-19 LOS and 1 on both COVID-19 mortality and LOS. Mortality was generally described as either in-hospital deaths, i.e., deaths occurring in a hospital or all-cause mortality, i.e., all deaths in COVID-19 patients, regardless of the cause. LOS was also defined commonly as the number of days in hospital admission due to COVID-19. One study described it as normal or prolonged, based on their measured average LOS (<17 days – normal; >17 - prolonged) (20). Consequently, they assessed LOS as a binary outcome. On the determinants of COVID-19 outcomes, identified risk factors were categorised into demographic, lifestyle, socioeconomic and biological/medical determinants, based on the determinants of health model (21). The rationale was to provide contextual analysis and identify common risk factors for COVID-19 outcomes. The biological/medical determinants encompassed as comorbidities, laboratory findings, and participants symptoms. The findings of the studies are synthesised below (Table 2).

**Table 2:**
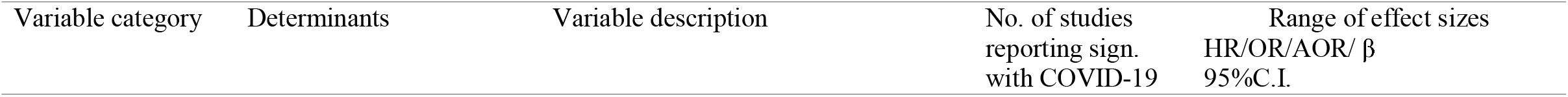
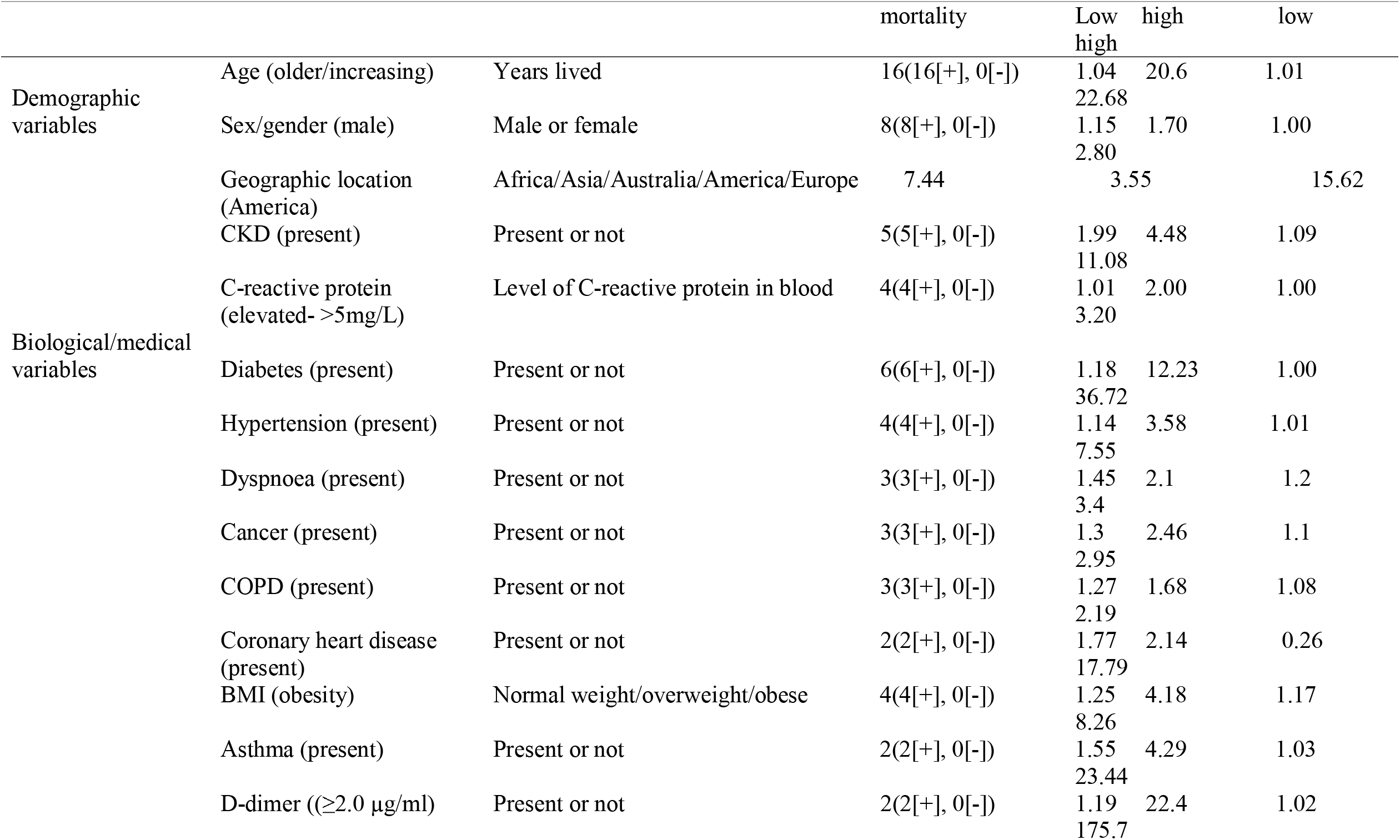
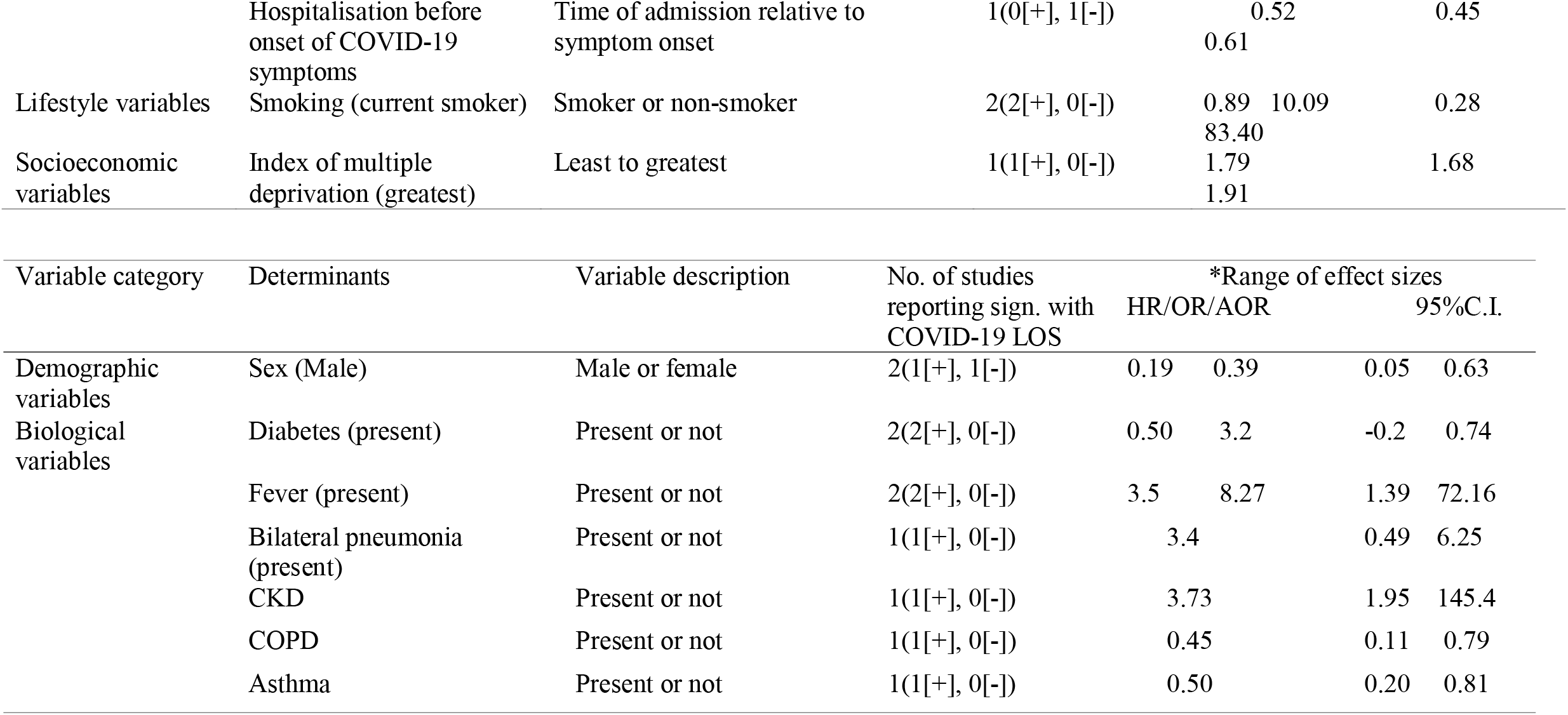
Synthesised review findings

Age and sex were the common demographic risk factors. Of the 20 studies on age and COVID-19 mortality, 16 identified increasing age as a significant determinant of COVID-19 mortality, with effect sizes ranging from 1.04 to 20.6 and 95% CI from 1.01 to 22.68 (9, 10, 11, 16, 17, 18, 24, 25, 26, 27, 28, 29, 30, 32, 33, 35). Also, 8 of the 20 studies on gender/sex and COVID-19 found men to have a higher risk of COVID-19 mortality than women (10, 11, 16, 17, 24, 26, 28, 32). On LOS, 2 of the 3 studies assessing LOS found women (AOR=0.19, 95%CI=0.05-0.63) (20) and men (β=0.39, 95% CI=0.16-0.62) (35) as determinants of COVID-19 duration of hospitalisation.

Of biological/medical risk factors, the review identified diabetes (n=6), Chronic kidney/renal disease (CKD) (n=5), hypertension (n=4), C-reactive protein (CRP) (n=4), BMI (n=4), dyspnoea (n=3), COPD (n=3), cancer (n=3), coronary heart disease (n=2), asthma (n=2) and D-dimer (n=2) as determinants of COVID-19 mortality. Of the 10 studies that included CKD in their analysis, 5 found it a significant determinant of COVID-19 mortality (18, 25, 26, 30, 35). Additionally, out of the 5 studies that investigated the influence of CRP on COVID-19 mortality, 4 showed that elevated CRP in the blood (at least >5mg/L) increases the risk of COVID-19 death (11, 24, 26, 29). Also, people with diabetes were found to have a higher risk of COVID-19 mortality in 6 of the 18 studies on diabetes and COVID-19 mortality (16, 17, 18, 26, 30, 32)

Like diabetes, hypertension was also identified as a determinant of COVID-19 mortality in 4 out of the 16 studies (11, 16, 24, 30). Furthermore, 4 of 11 studies on BMI and COVID-19 mortality showed that obesity significantly determines COVID-19 mortality (11, 18, 19, 32). For LOS, fever and diabetes were associated with prolonged LOS (43,35).

On lifestyle factors, smoking was the only assessed determinant of COVID-19 outcomes. It was identified as a risk factor for COVID-19 mortality in 2 of the eleven studies that explored it, with effect size ranging from 0.89-10.09, 95%CIs from 0.28-83.40 (23, 32). None of the studies on LOS reported a significant association between smoking and COVID-19 LOS. Finally, the only study on socioeconomic determinants and COVID-19 reported that greater deprivation determines COVID-19 mortality (HR=1.79, 95%1.68 – 1.91) (32)

## Discussion

This review aimed to investigate the determinants of COVID-19 outcomes. The review found that the specification and subsequent analysis of most of the determinants differed across the studies. For example, Berenguer et al. (2020) described elevated C-reactive protein (CRP) as CRP>5mg/L, while Carrasco-Sánchez et al. (2020) described it as >60mg/L. Also, whilst Almazeedi et al. (2020) and Sourij et al. (2020) assessed CRP as a continuous variable, Kaeuffer et al. (2020) categorised it into two groups: CRP-100-199mg/L and CRP≥200mg/L. Like CRP, older age was also specified differently across the studies. Bello-Chavolla et al. (2020) and Li et al. (2020) described it as individuals ≥65 years, Zhang et al. (2020) termed it as persons >65 years while Petrilli et al. (2020) defined it as people ≥75 years. Apart from these variations, the study settings also differed across the papers. These contextual differences, which could include disparities in access to healthcare, crowded living, could have inherently influenced the studies’ findings (36).

Despite these heterogeneities, the result of some of the determinants was similar across the studies. For instance, the significant association between age and COVID-19 mortality was reported across sixteen out of twenty papers, and four out of five articles examining CRP also showed a significant association between CRP and COVID-19 mortality. Regardless of the differences in methods, these findings could have public health implications for populations worldwide (37). Thus, they can be considered during the planning and implementation of effective policies for COVID-19.

Nonetheless, there were other contrasting findings. For example, Mendy et al. (2020) indicated that men are more likely to stay longer in hospitals due to COVID-19 than women. Conversely, Guo et al. (2020) showed that women are more associated with prolonged COVID-19 hospitalisation than men. Both studies had men dominating their study participants, 53% and 57% respectively; but the proportion of men in Guo et al. (2020) were marginally higher. However, in absolute figures, Mendy et al. (2020) included more male participants (n=365) than Guo et al. (2020) (n=43). Therefore, these sample size variations could account for the differences in their sex and LOS findings due to COVID-19. Moreover, these findings are from only two studies; thus, they may not be enough to conclude the association between sex and COVID-19 LOS. Further discussions on the review findings, based on the determinants of health model, are provided below.

### Demographic determinants of COVID-19 outcomes

The underlying mechanism for the association between older age and COVID-19 mortality is unclear; however, several studies indicate that decrease in immune responses coupled with increase comorbid burden with ageing may account for this observation (9, 38, 39). Another study further explained that age-related changes or defects in the immune system, particularly significant defect in cell-mediated immunity, primarily affects immune responses to diseases (40). Also, evolution and ageing theories, like the antagonistic pleiotropy theory, postulate that even beneficial genes at an early age may be less efficient or deleterious with increasing age, and this may inherently increase susceptibility to previously shielded diseases (41, 42). Moreover, current evidence suggests that increasing age is a common risk factor for several health outcomes, like mortalities and morbidities (43-46).

Even though ageing generally decreases immune responses to diseases and infections, the innate human response is mostly safeguarded (40). Thus, many individual and environmental factors may account for the relationship between ageing and disease outcomes, such as COVID-19 mortality. These factors may include nutritional deficiencies, decreased functionality, exposure to pathogens, vaccinations, an individual’s lived experiences and genetic make-up and access to health care (47). Furthermore, there are reports on good COVID-19 prognosis in elderly patients (48). Therefore, it may be imperative to understand how these factors cumulatively affect the immune system, and further mediate ageing and decreased immune system relationship to provide exhaustive literature on the subject. Other studies have also reported severe COVID-19 consequences in children (49, 50). Consequently, there is a need for studies to focus more on children, as much as they have on the adult population to offer a balanced argument to inform COVID-19 and ageing policies.

Like ageing, studies also attribute the sex disparities regarding COVID-19 mortality to sex-based differences in immunological responses to viral infections (51, 52). The X sex chromosome has encoded immune regulatory genes that decrease viral infections’ susceptibility (51). Since women have twice X-chromosomes to men, they tend to have higher innate immunity to viral infections, like COVID-19, and by extension a lower risk of severe COVID-19 outcomes than men (51, 53). Similarly, in contrast to oestrogen, testosterone has an immunosuppressive effect; so, it attenuates men’s immune responses to viral infections (54). Additionally, it is reported that men are genetically more predisposed to produce higher levels interleukin (IL)-6, which are unfavourable to longevity, compared to women (55). Apart from these biological reasons accounting for the sex differences in COVID-19 mortality risk, behavioural and lifestyle factors like smoking and alcohol consumption, have been implicated in the gender disparities in COVID-19 outcomes. Data indicate that men are more likely to engage in these lifestyle factors that increase the risk of COVID-19 deaths than women (52). Women are more likely to comply with COVID-19 precautionary measures than men and are more likely to remain confined than men (56). Regarding LOS, the evidence is not enough to indicate whether sex determines longer COVID-19 hospitalisation.

### Biological/medical determinants of COVID-19 outcomes

The biological/medical determinants in this review were comorbidities, symptoms, and laboratory findings of the included studies participants. The biological determinants of COVID-19 outcomes included CKD, C-reactive protein, diabetes, hypertension, obesity, cancer, COPD, dyspnea, asthma, and coronary heart disease. Clinical data reveal that chronic conditions, such as the above, decrease innate immune responses in humans (57, 58). For instance, metabolic diseases/disorders, like diabetes, attenuates immunity and increase risk to infections by weakening lymphocyte and macrophage activities (59). Moreover, these chronic conditions are associated with increase pro-inflammatory cytokines resulting from dysregulation of systems, like the hypothalamic-pituitary-adrenal and sympathetic nervous system (58, 60). The accumulation of pro-inflammatory cytokines subsequently impairs systemic and cellular immune functions (57, 61).

Furthermore, studies hypothesize that the use of renin-angiotensin-aldosterone system (RAAS) inhibitors, like angiotensin-converting enzyme-2 (ACE-2), in the management of some of these chronic conditions increase COVID-19 infectivity (62, 63). This is because ACE2 also functions as a receptor for the COVID-19 virus (64). This RAAS and ACE-2 hypothesis recently sparked debate and discourse on gold standard medical management of comorbidities in COVID-19 patients. One study indicated that sudden discontinuation of ACE-2 might have far worse consequences for high-risk-COVID-19 patients, particularly those with cardiovascular conditions, like myocardial infarction (65). Their argument is hinged on the paucity of human studies to corroborate the RAAS and ACE-2 theory. Additionally, experimental studies in mice showed that ACE-2 downregulation facilitates lung injuries and increases viral loads (60, 66). Thus, several human studies are needed to substantiate the harmful effect, or otherwise, of RAAS inhibitors in the management of COVID-19 patients with comorbidities.

### Lifestyle determinants of COVID-19 outcomes

The included studies examined only smoking as a lifestyle determinant of COVID-19 mortality. The association between smoking and COVID-19 mortality is biologically plausible because smoking is a risk factor for several conditions, like coronary heart disease and Chronic Obstructive Pulmonary Disease (COPD), that are associated with severe COVID-19 outcomes (67). Also, a cohort study with an average of 9.6 years follow-up by showed that 11% (men) and 13% (women) of pneumonia and COPD deaths were attributable to smoking (68). Additionally, the Centre for Disease Control and Prevention (CDC) also report that smoking is associated with about 113,000 respiratory deaths each year in the United States (69). Since COVID-19 is a respiratory infection and based on the above evidence on smoking-related respiratory deaths, it may be reasonable to make projections that smoking may be significantly associated with severe COVID-19 outcomes.

Furthermore, data shows that smokers have increased upregulation or expression of ACE-2, the reported enzyme receptor for SARS-CoV-2 (COVID-19) (70), which may increase their risk of severe COVID-19 outcomes compared to non-smokers. A single-cell sequencing experiment further demonstrated that cigarette smoking upregulates ACE-2 in humans’ lungs and increases their susceptibility to COVID-19 infections (71). They inferred that smoking cessation could reduce ACE-2 expression and thereby reduce the risk of COVID-19 disease. Thus, their findings advance the above argument on the benefit or otherwise of ACE-2 dysregulation in humans in reducing the burden of COVID-19. Consequently, systematic reviews and meta-analyses of several high-quality studies on ACE-2 and COVID-19 are required to provide empirical evidence to inform policy and clinical practice. All the same, this review is limited in drawing a meaningful conclusion on the association between smoking and COVID-19 mortality because only one of the included studies identified smoking as a risk factor of COVID-19 mortality.

### Policy implications

This review findings re-enforce the need for health systems to continue the testing, tracing, and isolation policies to reduce the spread of the virus and subsequently decrease the burden of the pandemic, particularly for high-risk individuals identified in this review. Additionally, countries, such as Nigeria, that are yet to implement crucial public health policies, like immunisation and routine COVID-19 testing, can draw lessons from countries like the UK that have benefited immensely from such policies. Although this may come with increased direct costs, especially the cost of PPEs and testing equipment, the indirect benefits to populations, such as reduced disease burden and improved productivity, might be enormous for these countries.

However, lockdown policies, which seems to be the go-to policy for most countries, must be evaluated holistically, to ascertain their overall benefits, especially as the pandemic continues to rage. Evidence shows that lockdowns are beneficial when introduced at an earlier stage of an outbreak than later. For example, evaluating the national lockdown response in Norway, USA, Argentina, and the UK shows that the lockdown’s timing and not the lockdown itself significantly reduced the burden of the outbreak in Argentina and Norway compared to the UK and the USA. For instance, the UK suffered significant health and economic recession due to the delays in the lockdown response to the viral outbreak. Their delay in implementing the first lockdown resulted in several other lockdowns that have had further economic implications. Therefore, lockdowns must be implemented earlier to prevent dire health and economic consequences.

Even the timing of lockdowns alone may not be enough to radically reduce viral transmission rate and consequently limit the probability of infections in high-risk individuals because data from other jurisdictions that introduced earlier national lockdowns, like Ghana, showed a steady increase in transmissions during the lockdown and a rapid rise few weeks post lockdown. Also, it is still crippling with the lockdown induced economic downfall. This suggests that lockdowns as single policies may be ineffective in plummeting COVID-19 infections, mortalities, and financial hardships. Consequently, global health systems must also place a premium on other equally important policies, like robust testing and tracing. Currently, most countries do not conduct follow-up COVID-19 testing following negative test results at entry borders. A negative test result on arrival at entry borders may not be enough to declare individuals as virus-free since it can take fourteen days maximum for the virus to be detected, per the WHO guidelines. Thus, governments must apply both on arrival and fourteen days post border arrival COVID-19 testing to ensure comprehensive identification of all positive cases to curtail the virus’s potential spread.

Additionally, it will be crucial for countries, like the USA and Brazil, to implement the fourteen-day quarantine policies for all border entries, as it has shown to be effective reducing the COVID-19 impact in places like Argentina. Countries with partial quarantine policies, such as Ghana, will benefit from instituting mandatory quarantine for all border arrivals at government selected facilities since self-isolation education and passenger locator forms may be inadequate in reducing the viral spread. Furthermore, health systems must continuously promote behavioural change interventions to establish significant control over the viral outbreak. Enforcing compliance to behavioural change interventions, like social distancing, nose mask use, social hand washing and cough etiquette, can be the significant catalyst needed to decrease the pandemic’s impact. Finally, the continuous increase in the COVID-19 cases and mortality indicate the need for an urgent review of current health policies at both international and national levels to implement suitable context-specific interventions to mitigate the COVID-19 menace effectively.

Concerning this study’s strength, this is the first systematic review synthesising the evidence on LOS determinants, to the best of the researchers’ knowledge. Therefore, it presents novel findings that could initiate useful interventions to address the burden of prolonged hospitalisation associated with COVID-19. It could also cause a paradigm shift and ensure holistic research coverage on all COVID-19 outcomes. Moreover, this is the first review to provide a comprehensive investigation of contextual determinants of COVID-19 outcomes, based on the determinants of health model. It identified crucial gaps in the literature on the determinants of COVID-19 outcomes that require urgent attention. Additionally, this review evaluated current public health policies and suggested strategies to augment area-specific efforts at curbing the COVID-19 problem. Regarding limitations, the review was restricted in conducting further analysis, specifically, meta-analysis, to precisely estimate the associations’ effect size due to included studies’ heterogeneity. Also, most of the included studies (n=12) used retrospective design, thus, there was the possibility of residual confounders that could influence this review’s findings. Additionally, all of them used secondary data from medical records of participants. Therefore, any omission or data entry error could affect their results and this review. Hence, caution must be taken when interpreting the findings of this review.

## Conclusion and recommendations

This study’s overarching aim was to examine the determinants of COVID-19 outcomes. The review findings showed that increasing age and comorbidities are more likely to determine COVID-19 outcomes. Thus, policies, like routine COVID-19 testing and prioritised vaccination to shield these high-risk individuals, must be sustained and extended to other populations yet to implement such important policies. Most importantly, health systems must continually review existing policies to ensure their context-specific relevance, especially with the emergence of a new viral variant and the rapid surge in cases. Based on this review, the authors recommend that future studies also focus on determinants of COVID-19 LOS. Additionally, studies should explore the determinants of COVID-19 outcomes in low-income-countries to ensure holistic and context-specific evidence on risk factors of COVID-19 mortality in the literature.

## Supporting information

PRISMA checklist

## Data Availability

All relevant data are either included in this article or added to the supplementary information.

## Contributors

SC and NKA conceptualised the research. SC and NKA conducted the systematic review with inputs from SP. SC wrote the first draft and NKA and SP revised the manuscript.

## Patient and public involvement

No direct involvement of patients or the public.

## Patient consent for publication

Not required.

## Data availability statement

All relevant data are either included in this article or added to the supplementary information.

