## Supplemental table 1 for "Determinants of COVID-19 outcomes: A systematic review"

Supplementary information 1: Full search strategy

| Database | Search terms | Date searched |
| --- | --- | --- |
| Scopus | (Determinant* OR Risk factor* OR Predictor*) AND (COVID-19 OR Coronavirus OR 2019 nCOV-2 OR SARS-COV-2) AND (Death OR Mortality*) AND (Length of hospitalisation stay OR Admission duration OR admission* length OR hospital* length). | 21^st^ December 2020 |
| Google Scholar | (Determinant* OR Risk factor* OR Predictor*) AND (COVID-19 OR Coronavirus OR 2019 nCOV-2 OR SARS-COV-2) AND (Death OR Mortality*) AND (Length of hospitalisation stay OR Admission duration OR admission* length OR hospital* length). | 23^rd^ December 2020 |
| CINAHL | (Determinant* OR Risk factor* OR Predictor*) AND (COVID-19 OR Coronavirus OR 2019 nCOV-2 OR SARS-COV-2) AND (Death OR Mortality*) AND (Length of hospitalisation stay OR Admission duration OR admission* length OR hospital* length). | 27^th^ December 2020 |
| Web of Science | (Determinant* OR Risk factor* OR Predictor*) AND (COVID-19 OR Coronavirus OR 2019 nCOV-2 OR SARS-COV-2) AND (Death OR Mortality*) AND (Length of hospitalisation stay OR Admission duration OR admission* length OR hospital* length). | 30^th^ – 31^st^ December 2020 |
