## Supplemental table 2 for "Determinants of COVID-19 outcomes: A systematic review"

Supplementary information 2: Data extraction questions

| *Themes* | *Review questions* |
| --- | --- |
| General Information | 1. Authors 2. Year 3. Aim 4. Country/Setting of study |
| Methodology | 5. What was the theoretical underpinning of the study? 6. What study design was used? 7. What sampling method was used? 8. What was the sample size? 9. What was the statistical basis of the sample size? 10. What variables were measured as determinants of COVID 19 mortality/length of hospital stay? 11. How were these variables specified? 12. How was data on these variables collected? 13. If primary data, what method was used to collect the data? 14. If secondary data, what dataset was used? 15. How was the outcome/dependent variable specified? 16. What statistical methods were used? 17. Were any statistical model diagnostics tests reported? 18. What are the main findings? 19. What were the author stated challenges? |
