## Supplemental table 3 for "Determinants of COVID-19 outcomes: A systematic review"

Supplementary Information 3: Quality and risk of bias assessment of the included studies.

Quality appraisal scores of the cohort studies ^a^

| Studies | Selection domain | | | | Comparability domain | Outcome domain | | | Total score |
| --- | --- | --- | --- | --- | --- | --- | --- | --- | --- |
|  | 1 | 2 | 3 | 4 | 1 | 1 | 2 | 3 |  |
| Alaa et al. (2020) | * | * | * | * | ** | * | * | * | 9 |
| Almazeedi et al. (2020) | * | * | * | * | ** | * | * | * | 9 |
| Berenguer et al. (2020) | * | * | * | * | * | * | * | * | 8 |
| Carrasco-Sánchez et al. (2020) | * | * | * | * | ** | * | * | * | 9 |
| Halalau et al. (2020) | * | * | * | * | * | * | * | * | 8 |
| Kaeuffer et al. (2020) | * | * | * | * | * | * | * | * | 8 |
| Li et al. (2020) | * | * | * | * | * | * | * | * | 8 |
| Okoh et al. (2020) | * | * | * | * | * | * | * | * | 8 |
| Petrilli et al. (2020) | * | * | * | * | ** | * | * | * | 9 |
| Sourij et al. (2020) | * | * | * | * | * | * | * | * | 8 |
| Wang et al. (2020) | * | * | * | * | * | * | * | * | 8 |
| Wu et al. (2020) | * | * | * | * | * | * |  | * | 7 |
| Zhou et al. (2020) | * | * | * | * | * | * |  | * | 7 |
| Tartof et al. (2020) | * | * | * | * | ** | * | * | * | 9 |
| Williamson et al. (2020) | * | * | * | * | ** | * | * | * | 9 |
| Grasselli et al. (2020) | * | * | * | * | * | * | * | * | 8 |
| Mikami et al. (2020) | * | * | * | * | ** | * | * | * | 9 |

*indicates study met the item.

^a^ Quality appraised with the Newcastle-Ottawa Quality Assessment Form for Cohort Studies (Peterson J, Welch V, Losos M, Tugwell PJ. The Newcastle-Ottawa scale (NOS) for assessing the quality of nonrandomised studies in meta-analyses. Ottawa: Ottawa Hospital Research Institute. 2011).

Quality appraisal scores of the cross-sectional studies ^b^

| Item/studies | Items | | | | | | | | | | |  |
| --- | --- | --- | --- | --- | --- | --- | --- | --- | --- | --- | --- | --- |
|  | 1 | 2 | 3 | 4 | 5 | 6 | 7 | 8 | 9 | 10 | 11 | Total |
| Bello-Chavolla et al. (2020) | Y | Y | Y | Y | N | N | NA | N | NA | Y | Y | 6 |
| Mendy et al. (2020) | Y | Y | Y | Y | N | N | Y | N | Y | Y | Y | 8 |
| Guo et al. (2020) | Y | Y | Y | Y | N | N | Y | N | Y | Y | Y | 8 |
| Albitar et al. (2020) | Y | Y | Y | Y | N | Y | Y | N | Y | N | Y | 8 |
| Zhang et al. (2020) | Y | Y | Y | Y | N | Y | NA | N | NA | Y | Y | 7 |

Y- yes, N- No, NA -not applicable.

^b^ Quality appraised with the Agency for Healthcare Research and Quality (ARHQ) Checklist for Cross sectional studies (Zeng X, Zhang Y, Kwong JS, Zhang C, Li S, Sun F, Niu Y, Du L. The methodological quality assessment tools for preclinical and clinical studies, systematic review and meta‐analysis, and clinical practice guideline: a systematic review. Journal of evidence-based medicine. 2015 Feb;8(1):2-10).

Risk of bias assessment

| Studies | Domain | | | | | |
| --- | --- | --- | --- | --- | --- | --- |
|  | Selection of participants | Confounding | Exposure measurement | Outcome assessment | Study attrition | Study outcome reporting |
| Alaa et al. (2020) | Low risk | Low risk | Low risk | Low risk | Low risk | Low risk |
| Almazeedi et al. (2020) | Low risk | Low risk | Low risk | Low risk | Low risk | Low risk |
| Bello-Chavolla et al. (2020) | Low risk | moderate risk | Low risk | Low risk | Low risk | Low risk |
| Berenguer et al. (2020) | Low risk | moderate risk | Low risk | Low risk | Low risk | Low risk |
| Carrasco-Sánchez et al. (2020) | Low risk | Low risk | Low risk | Low risk | Low risk | Low risk |
| Halalau et al. (2020) | Low risk | moderate risk | Low risk | Low risk | Low risk | Low risk |
| Kaeuffer et al. (2020) | Low risk | Moderate risk | Low risk | Low risk | Low risk | Low risk |
| Li et al. (2020) | Low risk | moderate risk | Low risk | Low risk | Low risk | Low risk |
| Okoh et al. (2020) | Low risk | moderate risk | Low risk | Low risk | Low risk | Low risk |
| Petrilli et al. (2020) | Low risk | Low risk | Low risk | Low risk | Low risk | Low risk |
| Sourij et al. (2020) | Low risk | Moderate risk | Low risk | Low risk | Low risk | Low risk |
| Wang et al. (2020) | Low risk | Moderate risk | Low risk | Low risk | Low risk | Low risk |
| Zhang et al. (2020) | Low risk | Moderate risk | Low risk | Low risk | Low risk | Low risk |
| Zhou et al. (2020) | Low risk | Moderate risk | Low risk | Low risk | Low risk | Low risk |
| Tartof et al. (2020) | Low risk | Low risk | Low risk | Low risk | Low risk | Low risk |
| Williamson et al. (2020) | Low risk | Low risk | Low risk | Low risk | Low risk | Low risk |
| Grasselli et al. (2020) | Low risk | Moderate risk | Low risk | Low risk | Low risk | Low risk |
| Mikami et al. (2020) | Low risk | Low risk | Low risk | Low risk | Low risk | Low risk |
| Albitar et al. (2020) | Low risk | Moderate risk | Low risk | Low risk | Moderate risk | Low risk |
| Wu et al. (2020) | Low risk | Moderate risk | Low risk | Moderate risk | Low risk | Low risk |
| Guo et al. (2020) | Low risk | Moderate risk | Low risk | Low risk | Low risk | Low risk |
| Mendy et al. (2020) | Low risk | Moderate risk | Low risk | Low risk | Low risk | Low risk |

Risk assessed with the risk of bias assessment tool for non-randomised studies (RoBANS). Park J, Lee Y, Seo H, Jang B, Son H, Kim S, Shin S, Hahn S. Risk of Bias Assessment tool for Non-randomized Studies (RoBANS): Development and validation of a new instrument. In: Abstracts of the 19th Cochrane Colloquium; 2011 19-22 Oct; Madrid, Spain. John Wiley & Sons; 2011.
